# Frailty outweighs psychiatric variables in predicting clinical outcomes among older adults receiving consultation-liaison psychiatry: a multicentre prospective cohort study (OLD-3 Study)

**DOI:** 10.64898/2026.06.29.26356821

**Authors:** Leire Narvaiza-Grau, Miguel Alonso-Sanchez, Jorge Cuevas-Esteban, Monica Prat-Galbany, Eduardo Delgado-Parada, Cristina Pujol-Riera, Beatriz Villagrasa-Blasco, Sara Crivilles-Mas, Mikel Etxandi-Santolaya, Nestor Arbelo-Cabrera, Paloma Muñoz-Calero, Iñigo Alberdi-Paramo, Mar Baz, Sara Lakis-Granell, Eduardo Fuster-Nacher, Maria Iglesias-Gonzalez, The Liaison Geriatric Psychiatry Working Group of the Spanish Psychogeriatric Society

## Abstract

**Background:** Older adults evaluated by consultation-liaison psychiatry services (CLPS) often present with complex psychiatric and medical comorbidity, frequent psychotropic exposure and a high prevalence of frailty. However, the relative contribution of these factors to clinical outcomes remains unclear.

**Methods:** We conducted a multicentre prospective cohort study including 465 consecutive patients aged ≥65 years evaluated by CLPS in 10 general hospitals in Spain between January and July 2024. Psychiatric history, post-consultation psychiatric diagnoses, psychotropic use, age group (65–74 vs ≥75 years) and frailty assessed using the Clinical Frailty Scale were recorded. Outcomes included falls, institutionalisation, access to mental health follow-up and mortality at 1 and 3 months after discharge.

**Results:** The mean (SD) age was 77.4 (7.8) years and 55.9% were women. Psychiatric history was present in 68.8% of patients and 55.8% received a new psychiatric diagnosis, most commonly delirium. Psychotropic use was frequent (71.6%), particularly antidepressants (49.0%) and benzodiazepines (42.6%). Psychotropic polypharmacy was associated with falls. Frailty was prevalent (60.2%) and independently associated with early mortality, whereas age was the main predictor of mortality between 1 and 3 months. Older age was also associated with a lower likelihood of specialised mental health follow-up.

**Conclusions:** Among older adults evaluated by CLPS, frailty and medical comorbidity appear to outweigh psychiatric variables in predicting outcomes. Integrating comprehensive geriatric assessment and medication review into CLPS may improve care for this population.

## INTRODUCTION

Population aging has been associated with a sustained rise in hospitalizations among older adults, who frequently present with high levels of medical and psychiatric comorbidity. This complexity increases the risk of complications, functional decline, and mortality and requires specialized, integrated care. In this context, consultation-liaison psychiatry services play a key role in diagnostic assessment and the optimization of psychotropic treatment, particularly in geriatric populations (1,2). In addition, integrated consultation-liaison psychiatry models have been shown to be cost-effective, supporting their implementation in general hospitals (1).

Among hospitalized older adults, the most common psychiatric conditions include delirium, depression, and dementia (3), all of which are associated with functional decline, institutionalization, antipsychotic use, and increased mortality. The use of psychotropic medications increases with age; in particular, benzodiazepines and antipsychotics have been associated with a higher risk of falls, fractures, and morbidity and mortality (4–6). Furthermore, these medications are often prescribed outside recommended guidelines or continued longer than advised, which may contribute to poorer outcomes, including readmissions and mortality (7).

Frailty, which is highly prevalent among older adults with mental disorders, is independently associated with mortality, falls, institutionalization, and disability, although evidence in the specific context of consultation-liaison psychiatry remains limited (8). Clinically relevant differences have also been described between patients aged 65–74 years and those aged 75 years and older, with potential prognostic and therapeutic implications (3,9,10).

Despite these findings, evidence remains limited regarding the clinical characteristics of these patients, the prevalence of frailty, patterns of psychotropic prescribing, and potential disparities in access to consultation-liaison psychiatry services. Agerelated barriers to mental health care have been suggested; however, empirical evidence supporting these disparities remains scarce (11). In this context, the Geriatric Consultation-Liaison Psychiatry Working Group of the Spanish Psychogeriatric Society (SEPG) conducted the The Old Liaison Dementia-Delirium-Depression study (OLD-3). The primary objective was to describe the sociodemographic and clinical characteristics of patients aged 65 years and older referred to consultation-liaison psychiatry. Secondary objectives were to (1) identify high-risk psychotropic medications and their association with falls; (2) evaluate clinical differences and risk of institutionalization between patients aged 75 years and older and those aged 65–74 years; (3) explore potential age-related disparities in access to care; and (4) assess the prevalence of frailty and its associated factors.

## METHODS

### Study Design and Participants

This study was conducted within the Liaison Geriatric Psychiatry Working Group of the Spanish Psychogeriatric Society, a collaborative network of psychiatrists with expertise in geriatric psychiatry and consultation-liaison psychiatry for older adults. Project OLD-3 is one of the research initiatives developed within this network and involved 10 general hospitals across Spain. All participating investigators were psychiatrists with clinical expertise in the mental health care of older adults.

The OLD-3 study was a prospective, multicenter observational study in which patients were consecutively recruited from consultation-liaison psychiatry services at the participating hospitals (Supplement 1). The study consisted of two phases: recruitment during hospital admission and post discharge follow-up through review of electronic health records.

The inclusion period extended from January 15 to July 15, 2024. Eligible participants were patients aged 65 years and older referred to consultation-liaison psychiatry from medical or surgical departments of general hospitals who provided written informed consent.

Exclusion criteria included patients referred exclusively to psychology services, those evaluated in emergency departments, those treated in psychogeriatric units, those enrolled in specific proactive intervention programs, patients unable to provide consent without a legal representative, and those who declined participation.

### Variables and Measures

Sociodemographic variables included age, sex, living situation, marital status, and educational level. Clinical variables included functional status, comorbidity, and frailty. Functional status was assessed using the Barthel Index for basic activities of daily living and the Lawton and Brody Scale for instrumental activities. Comorbidity was quantified using the age-adjusted Charlson Comorbidity Index.

Frailty was assessed using the Clinical Frailty Scale (CFS), a 9-point clinical scale that classifies vulnerability from very fit to terminally ill (12–15).

Additional variables included reason for hospital admission, reason for psychiatric consultation, psychiatric history, prior mental health care, psychiatric diagnosis following consultation, discharge destination, and history of falls within the 6 months prior to admission.

Perceived loneliness was assessed using the validated Spanish version of the University of California, Los Angeles Loneliness Scale (UCLA), specifically the 3-item version (Three-Item Loneliness Scale [TILS]) designed for rapid screening (16). These scales were administered only when patients were able to provide valid selfreport; otherwise, they were not applied.

Pharmacological variables included psychotropic treatment prior to admission, changes during hospitalization (initiation, discontinuation or dose adjustment), and psychotropic treatment at discharge following consultation-liaison psychiatry intervention.

### Clinical Procedures

Clinical evaluations were conducted by consultation-liaison psychiatry teams as part of routine clinical care during hospitalization. The frequency and number of assessments were determined according to clinical judgment based on patient needs.

Psychiatric diagnoses were established according to the Diagnostic and Statistical Manual of Mental Disorders, Fifth Edition, Text Revision (DSM-5-TR) by consultation-liaison psychiatrists (attending physicians and/or supervised residents), based on clinical interview, mental status examination, information from relatives or caregivers, and review of medical records. Diagnostic comorbidity was allowed; the primary diagnosis was defined as the main reason for consultation.

### Follow-up and Outcomes

Follow-up assessments were conducted at 1 and 3 months after hospital discharge through review of electronic health records.

At 1 month, the following outcomes were recorded: all-cause mortality, emergency department visits, hospital readmissions, and psychotropic prescriptions obtained through the electronic prescribing system.

At 3 months, mortality and psychotropic treatment were reassessed.

### Data Management

Each participating center independently collected data and reviewed medical records of patients within its catchment area. Data were subsequently anonymized and stored in a centralized database using REDCap.

### Statistical Analysis

Continuous variables were summarized as mean (SD) or median (IQR), as appropriate. Categorical variables were expressed as absolute frequencies and percentages.

Comparisons between categorical variables were performed using the χ^2^ test or Fisher exact test, as appropriate. Comparisons between continuous variables were conducted using the Student t test or the Mann–Whitney U test. For comparisons involving more than 2 groups, analysis of variance (ANOVA) or the Kruskal–Wallis test was used.

Binary logistic regression models were used to identify factors independently associated with key clinical outcomes, including discharge disposition, hospital readmissions, emergency department visits, and in-hospital mortality at 1 and 3 months. Covariates were selected based on prior evidence from the scientific literature, including age and sex, and on their significance in the bivariate analysis, according to the prespecified level of statistical significance (P⍰<⍰.05).

Statistical significance was set at a 2-sided P < .05. No adjustments for multiple comparisons were applied given the exploratory nature of the study.

All analyses were performed using IBM SPSS Statistics, version 29.0 (IBM Corp).

### Ethical Considerations

The study was approved by the Research Ethics Committee of Hospital Universitari de Bellvitge (Ref. PR069/23 [CSI 23/13]) and by the appropriate regulatory authorities at all participating centers. The study was conducted in accordance with the Declaration of Helsinki and applicable clinical research regulations (8.1,9.1).

Written informed consent was obtained from all participants or their legal representatives. The study also complied with European data protection regulations.

### Use of artificial intelligence tools

ChatGPT (OpenAI) was used to assist in improving the English language of the manuscript. The final version was subsequently reviewed and edited by several authors to ensure accuracy and clarity.

## RESULTS

### 1. Sociodemographic Characteristics

The sample included 465 patients; the characteristics of the sample are detailed in **Table 1**. The mean length of hospital stay was 23.85 days (SD, 23.60), with a median of 16 days (IQR, 9–29) and a range of 0 to 196 days (n = 455). During hospitalization, patients received a mean of 3.02 consultation-liaison psychiatry visits (SD, 2.52), with a median of 2 visits (IQR, 1–4) and a range of 1 to 27.

**Table 1.**
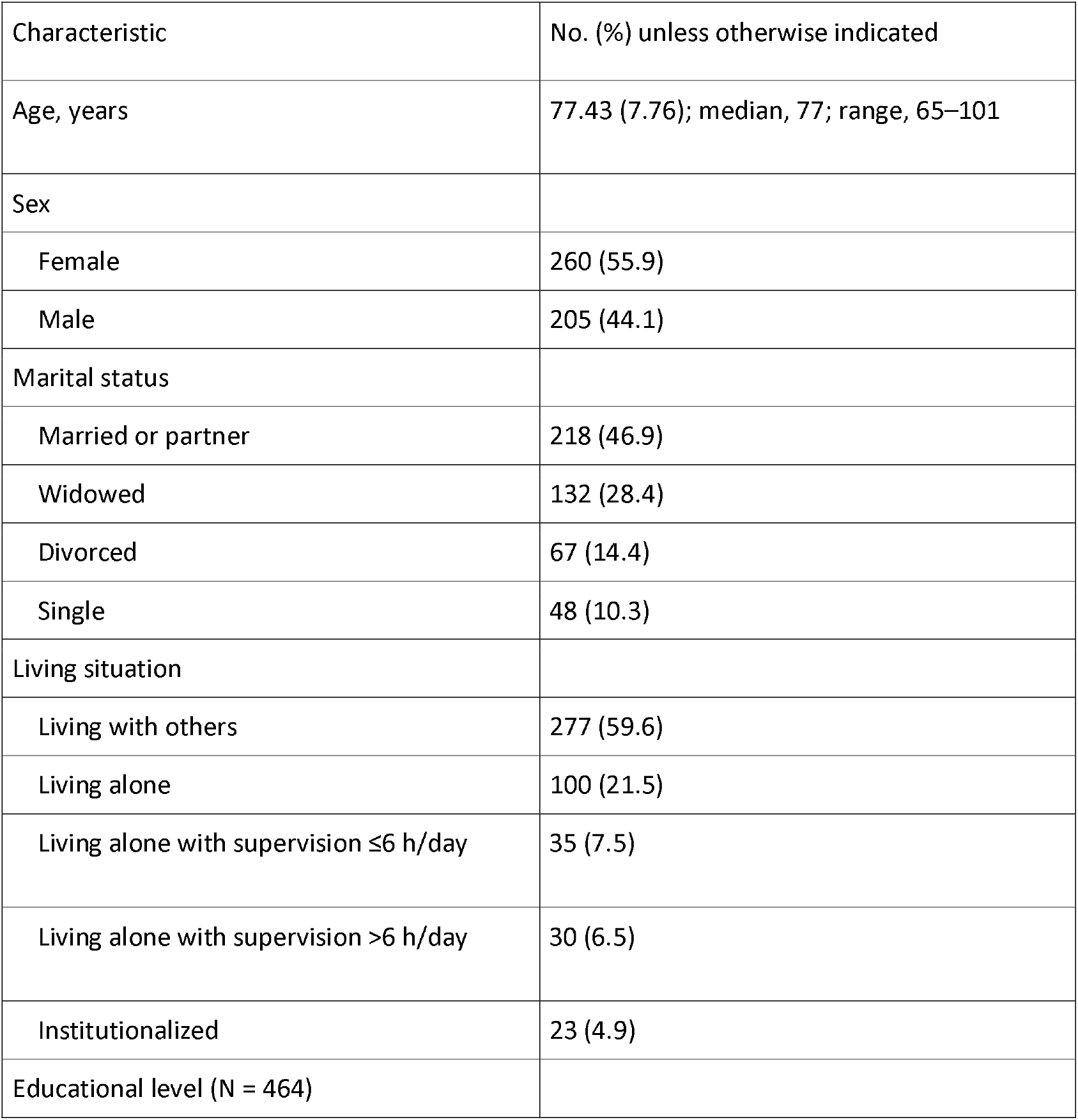
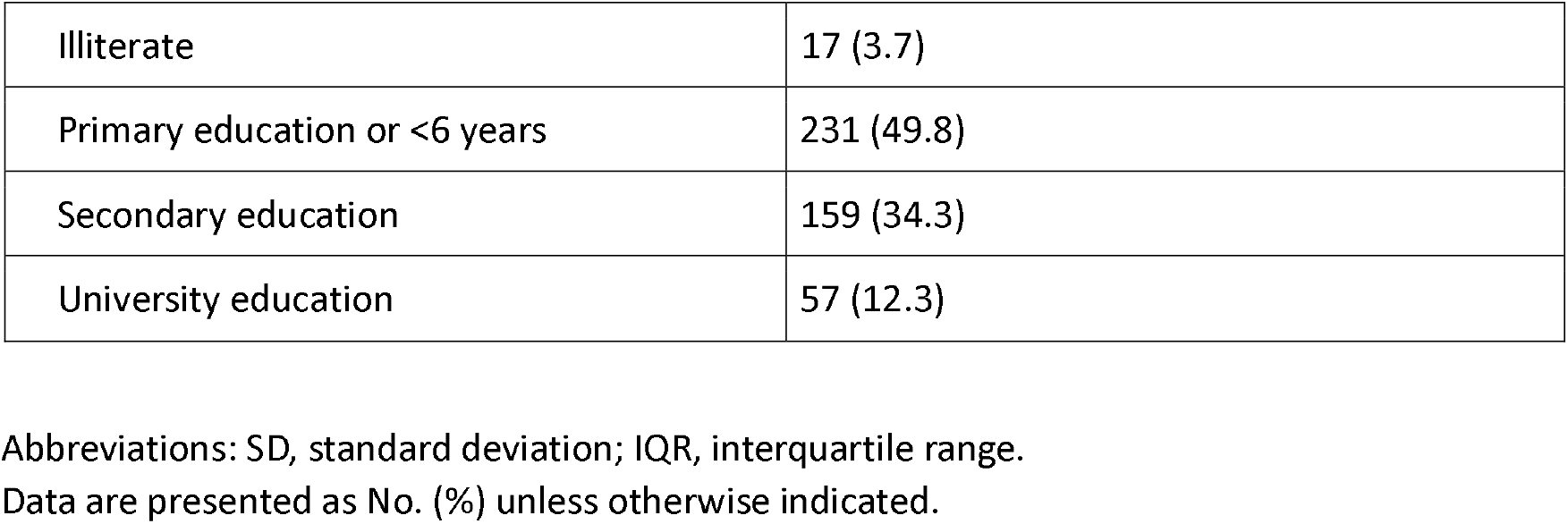
Sociodemographic Characteristics of the Study Sample.

### 2. Clinical Characteristics

Functional status: Baseline and admission functional status (Barthel Index), comorbidity (Charlson Index), dependence in instrumental activities (Lawton Index), as well as mobility prior to and at admission are shown in **Table 2**.

**Table 2.** Clinical Characteristics of the Study Sample.

| Characteristic | No. (%) unless otherwise indicated |
| --- | --- |
| Function, comorbidity, frailty, and mobility |  |
| Barthel Index prior to admission (N = 464) | Mean (SD), 81.6 (24.2); median (IQR), 90 (70–100); range, 0–100 |
| Barthel categories prior to admission |  |
| Total dependence | 14 (3.0) |
| Severe dependence | 83 (17.9) |
| Moderate dependence | 140 (30.2) |
| Mild dependence | 25 (5.4) |
| Independence | 202 (43.5) |
| Barthel Index at admission (N = 463) | Mean (SD), 55.4 (30.9); median (IQR), 60 (30–85); range, 0–100 |
| Barthel categories at admission |  |
| Total dependence | 89 (19.2) |
| Severe dependence | 177 (38.2) |
| Moderate dependence | 133 (28.7) |
| Mild dependence | 12 (2.6) |
| Independence | 52 (11.2) |
| Age-adjusted Charlson Comorbidity Index (N = 455) | Mean (SD), 5.41 (2.46); median (IQR), 5 (4–7); range, 0–14; ≥5 points: 271 (59.6) |
| Lawton–Brody Index (N = 458) | Median (IQR), 5 (3–8); moderate–severe dependence ($\leq 4$): 204 (44.5); functional independence ($\geq 7$): 140 (30.6) |
| Clinical Frailty Scale (CFS) (N = 460) | Median (IQR), 4 (3–6); non-frail (1–3): 183 (39.8); frail ($\geq 4$): 277 (60.2); moderate–severe ($\geq 5$): 170 (37.0) |
| Falls in the last 6 months (N = 464) | No: 332 (71.6); Yes: 132 (28.4) |
| Mobility prior to admission (N = 465) | Independent: 256 (55.1); cane: 94 (20.2); walker: 76 (16.3); dependent: 39 (8.4) |
| Mobility at admission (N = 465) | Independent: 142 (30.5); cane: 49 (10.5); walker: 70 (15.1); dependent: 204 (43.9) |
| Perceived loneliness |  |
| Perceived loneliness according to TILS (N = 409) |  |
| Significant loneliness ($\geq 6$ points) | 97 (23.7) |
| No loneliness | 312 (76.3) |
| TILS score (3 items; N = 442) | Median, 4; range, 0–9 |

Abbreviations: SD, standard deviation; IQR, interquartile range; CFS, Clinical Frailty Scale; TILS, Three-Item Loneliness Scale.

#### Falls

A total of 28.5% of patients (133 out of 465) experienced at least one fall in the 6 months prior to admission. In the bivariate analysis, patients with falls showed worse baseline functional status, with significantly lower scores on the Barthel Index (78.6 ± 24.7 vs 82.9 ± 23.8; p < 0.001) and the Lawton–Brody Index (3.98 ± 2.73 vs 4.86 ± 2.68; p = 0.002), compared with those without falls. They also had higher levels of perceived loneliness, with higher scores on the TILS scale (4.44 ± 2.41 vs 3.86 ± 1.90; p = 0.019).

From a clinical perspective, no statistically significant association was observed between the presence of psychiatric history and falls in the previous 6 months (30.9% vs 22.9%; χ^2^ = 2.76; p = 0.097; odds ratio [OR], 1.51; 95% CI, 0.96–2.38). Similarly, the type of primary psychiatric history was not significantly associated with the presence of falls (χ^2^ = 13.52; p = 0.26). However, falls were more frequent in patients with a post-consultation diagnosis of depressive disorder (43.8%; p < 0.05), delirium (40.7%; p < 0.05), and psychotic disorders (40.9%; p < 0.05).

Regarding prior pharmacological treatment, the use of antidepressants was significantly more frequent among patients with falls (59.4% vs 46.6%; p = 0.019), as was the use of mood stabilizers (19.6% vs 11.7%; p = 0.043). No significant differences were observed in prior use of benzodiazepines (43.8% vs 43.6%) or antipsychotics (28.1% vs 21.8%).

Psychotropic polypharmacy (≥2 classes) was more frequent in the group with falls (47.0% vs 35.6%; p = 0.032). In contrast, no significant associations were observed with age (p = 0.248) or with frailty assessed using the CFS (p = 0.185). In the multivariate analysis, neither age, frailty, nor functional indices remained independently associated with the presence of falls.

#### Frailty

Frailty was common in the sample. Among a total of 460 evaluated patients, the median Clinical Frailty Scale (CFS) score was 4 (IQR, 3–6). A total of 60.2% of patients (n = 277) were classified as frail (CFS ≥4), whereas 39.8% (n = 183) were considered non-frail (CFS 1–3). In addition, 37.0% of the sample (n = 170) presented moderate to severe frailty (CFS ≥5).

In the analysis by pharmacological groups prior to admission, only prior use of antipsychotics was significantly associated with greater frailty as measured by the CFS. Patients exposed to antipsychotics (n = 107) had a mean CFS score of 4.86, compared with 3.96 in those not exposed (p < 0.001). No significant differences in CFS scores were observed between patients with and without prior use of antidepressants (n = 228; 4.27 vs 4.07; p = 0.153), benzodiazepines (n = 198; 4.06 vs 4.25; p = 0.166), or mood stabilizers (n = 63; 4.27 vs 4.15; p = 0.813). Similarly, frailty did not differ significantly according to the presence of psychiatric history (4.25 vs 4.01; p = 0.156) or prior mental health follow-up (4.27 vs 4.14; p = 0.322)

#### Sensory Deficits

Among the 464 evaluated participants, 40.3% (n = 187) had visual impairment. Among patients with visual impairment, most used corrective glasses (89.3%; n = 167).

Regarding hearing, among a total of 463 participants, 22.0% (n = 102) had hearing loss. Among patients with hearing loss, only 25.5% (n = 26) used hearing aids.

#### Substance Use

A total of 24.5% of participants (n = 114) reported use of any substance in the previous year, with alcohol and tobacco being the most frequent; detailed results are presented in Table 3.

**Table 3.**
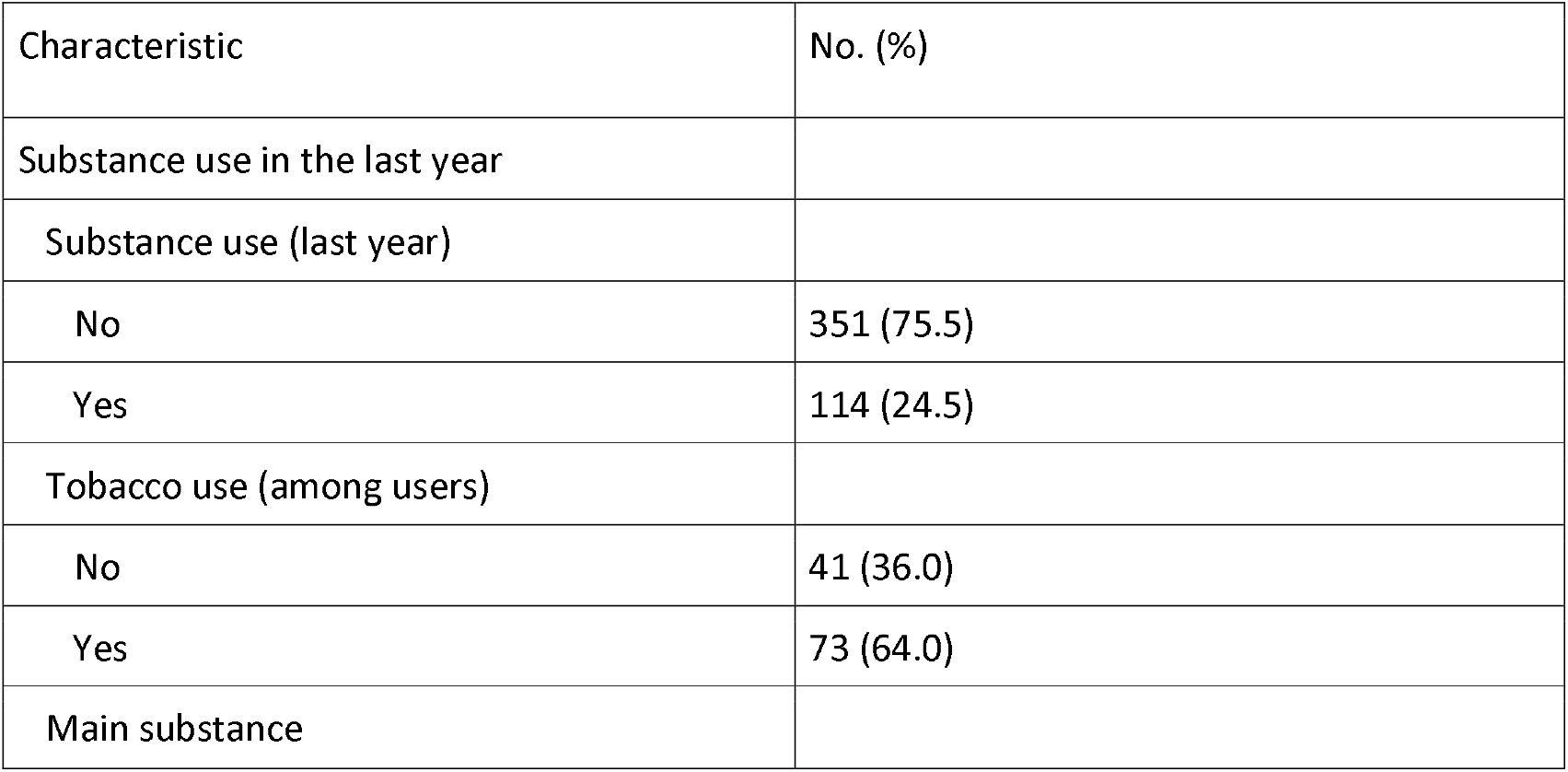
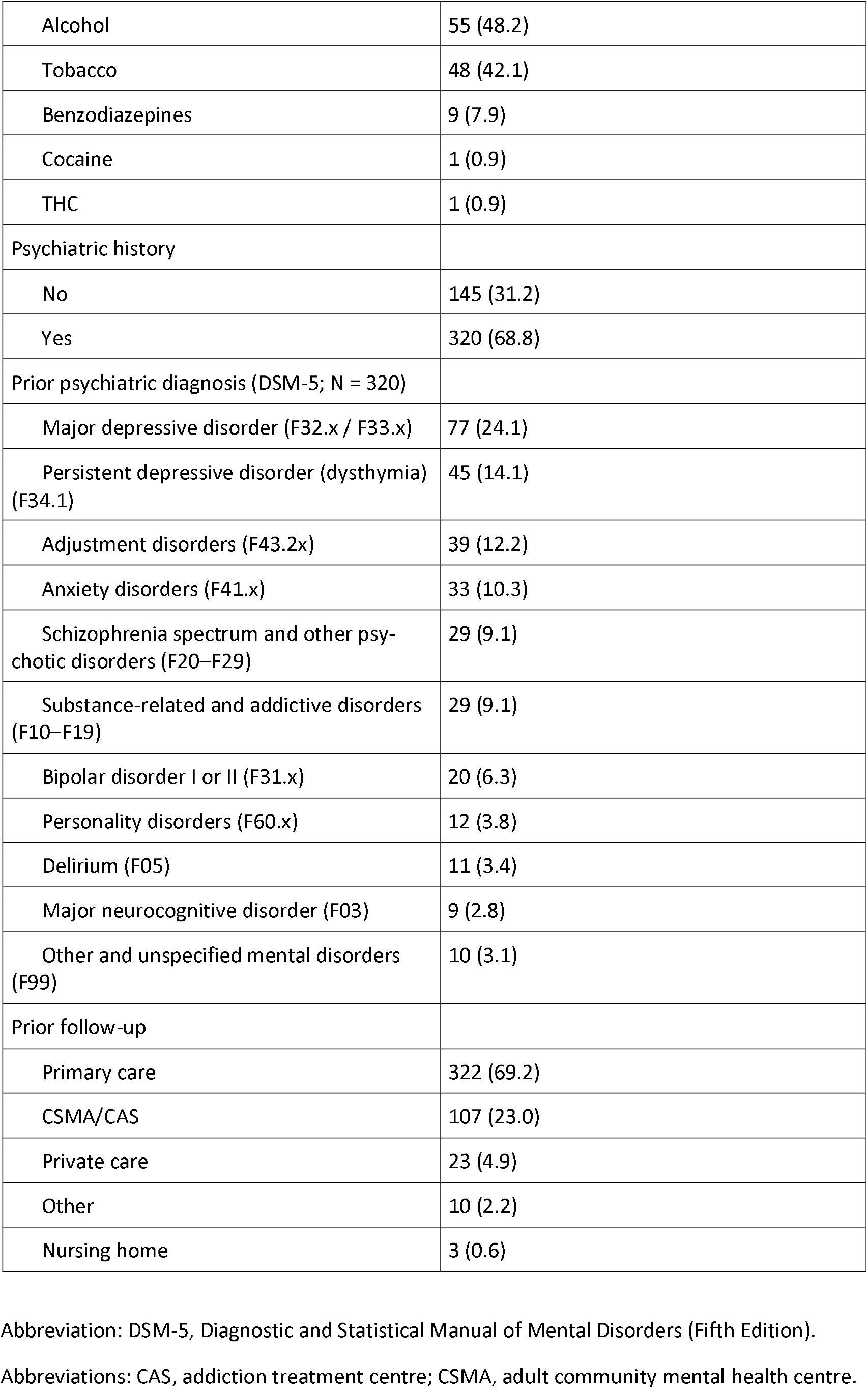
Substance Use, Psychiatric History, and Prior Follow-up.

#### Psychiatric History

A total of 68.8% (n = 320) of patients had a psychiatric history, with mood disorders being the most frequent.

Regarding suicidal behavior, 11.0% (n = 51) of the sample had a history of suicide attempts, and 23.5% (n = 12) had made a suicide attempt in the 12 months prior.

Regarding follow-up prior to admission, most patients were managed in primary care (69.2%, n = 322), followed by community mental health services (CSMA/CAS) (23.0%, n = 107), private care (4.9%, n = 23), and other resources (2.2%). Patients with psychiatric history who were receiving specialized follow-up accounted for 33.1% (n = 106).

In the multivariable logistic regression model, adjusted for sex, age-adjusted Charlson Comorbidity Index, Barthel Index, Lawton–Brody Index, and Clinical Frailty Scale, age was inversely associated with the likelihood of prior follow-up in CSMA/CAS (odds ratio [OR] per year, 0.95; 95% CI, 0.92–0.99; p = .006). None of the other variables included in the model showed a statistically significant association.

#### Prior Psychopharmacological Treatments

Among the 465 patients included in the cohort, 333 (71.6%) were receiving at least one psychotropic medication prior to admission. Antidepressants were the most frequently prescribed group (228 [49.0%]), followed by benzodiazepines (198 [42.6%]), antipsychotics (107 [23.0%]), and mood stabilizers (63 [13.5%]); other psychotropic medications were recorded in 21 patients (4.5%).

Among antidepressant users, the most commonly prescribed agents were sertraline (21.9%), venlafaxine (16.2%), duloxetine (9.6%), and mirtazapine (9.6%), followed by citalopram (8.3%), trazodone (7.0%), and paroxetine (5.7%). Concomitant treatment with ≥2 antidepressants was observed in 17.0% of the total sample and with ≥3 in 1.5%.

Among benzodiazepine users, lorazepam was the most frequently prescribed drug (41.4%), followed by alprazolam (14.6%), lormetazepam (14.6%), and diazepam (12.1%); 6.5% of patients received ≥2 benzodiazepines and 0.9% ≥3.

Quetiapine was the most commonly prescribed antipsychotic (58.9%), followed by risperidone (12.1%), olanzapine (10.3%), and paliperidone (6.5%). Overall, 4.5% of patients received ≥2 antipsychotics and 0.6% ≥3.

Among patients receiving mood stabilizers, the most frequent agents were pregabalin (38.1%), gabapentin (23.8%), lithium (19.0%), and valproate (14.3%); 1.1% of the total sample received ≥2 drugs in this group.

When stratified by psychiatric history, psychotropic use was higher across all pharmacological groups among patients with documented psychiatric history. In contrast, among those without psychiatric history, benzodiazepines were the most frequently prescribed psychotropic medications (43 [29.7%]), followed by antidepressants (13 [9.0%]), whereas the use of antipsychotics, mood stabilizers, and other psychotropic medications was very low (≤4 patients; ≤2.8%).

### 3. Consultation-Liaison Psychiatry Service Activity, Psychiatric Diagnoses, and Disposition

The location of evaluation, primary medical diagnosis, and reason for consultation are described in Table 4. After consultation, 55.8% of patients received a new psychiatric diagnosis. Among patients with complete data on the psychiatric diagnosis established after consultation (N = 426), the most frequent diagnosis was delirium, present in 108 patients (25.4%), followed by adjustment or trauma-related disorder in 102 patients (23.9%) and depressive disorder in 48 patients (11.3%). In 127 cases, more than one psychiatric diagnosis was made after consultation.

**Table 4.**
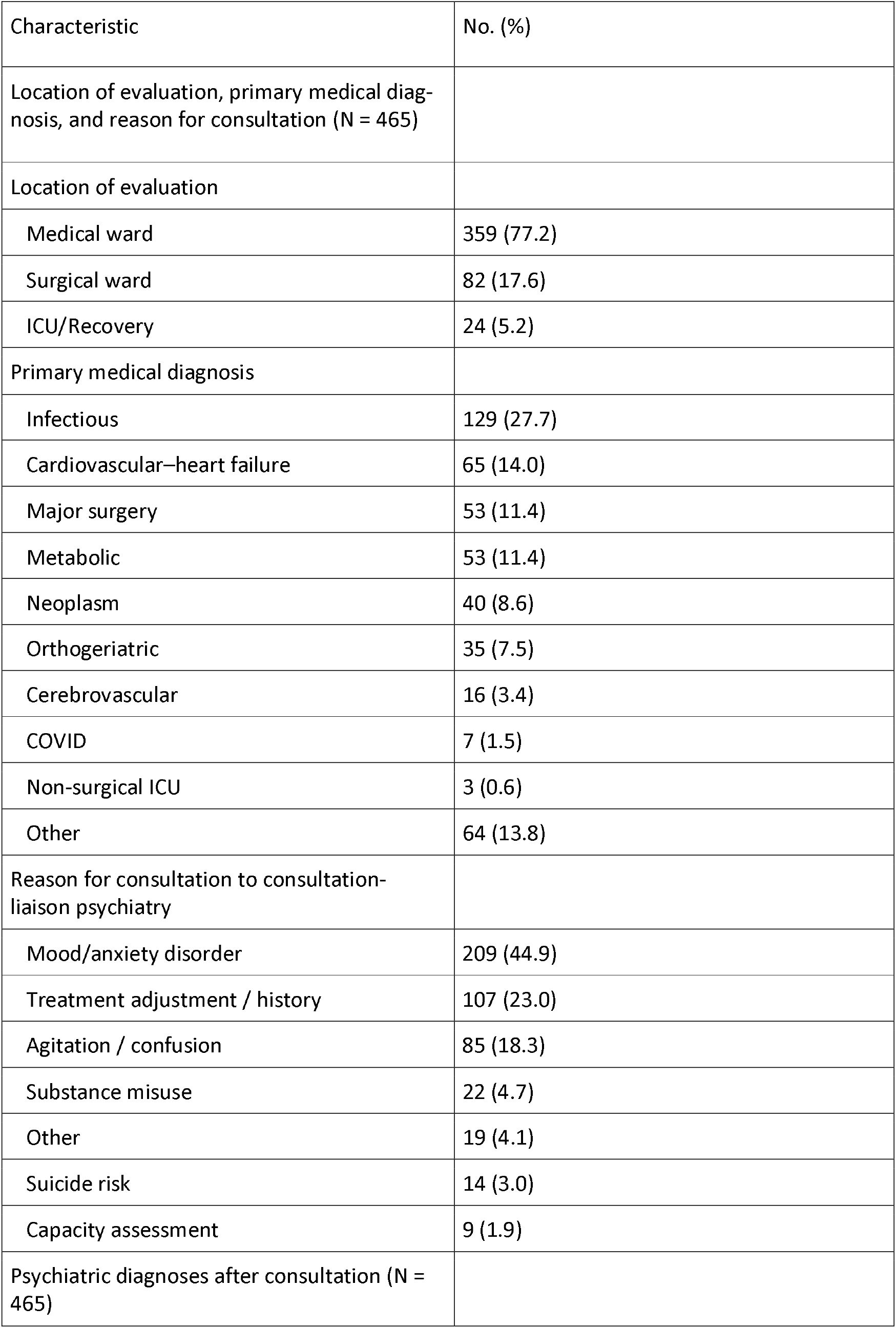
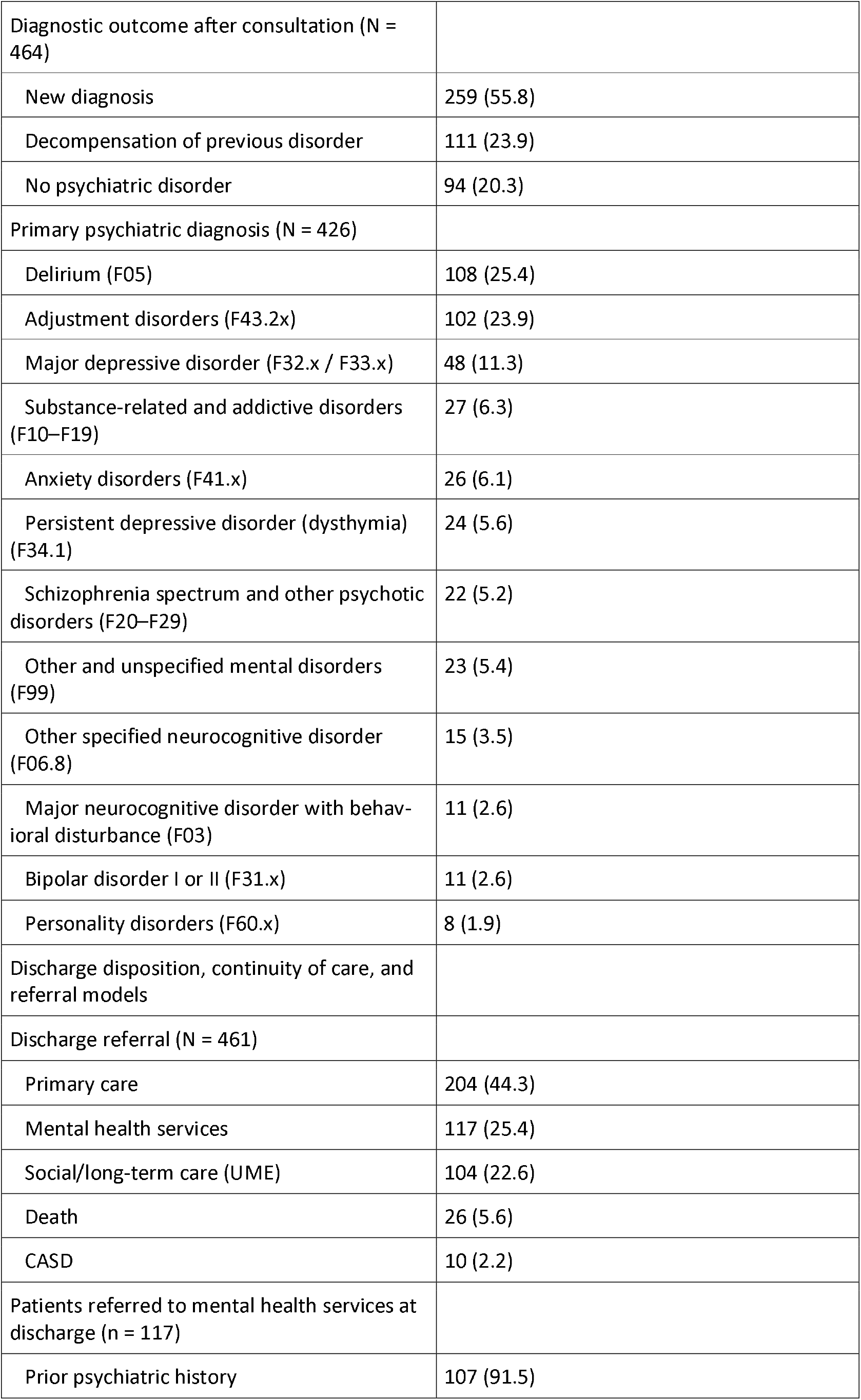
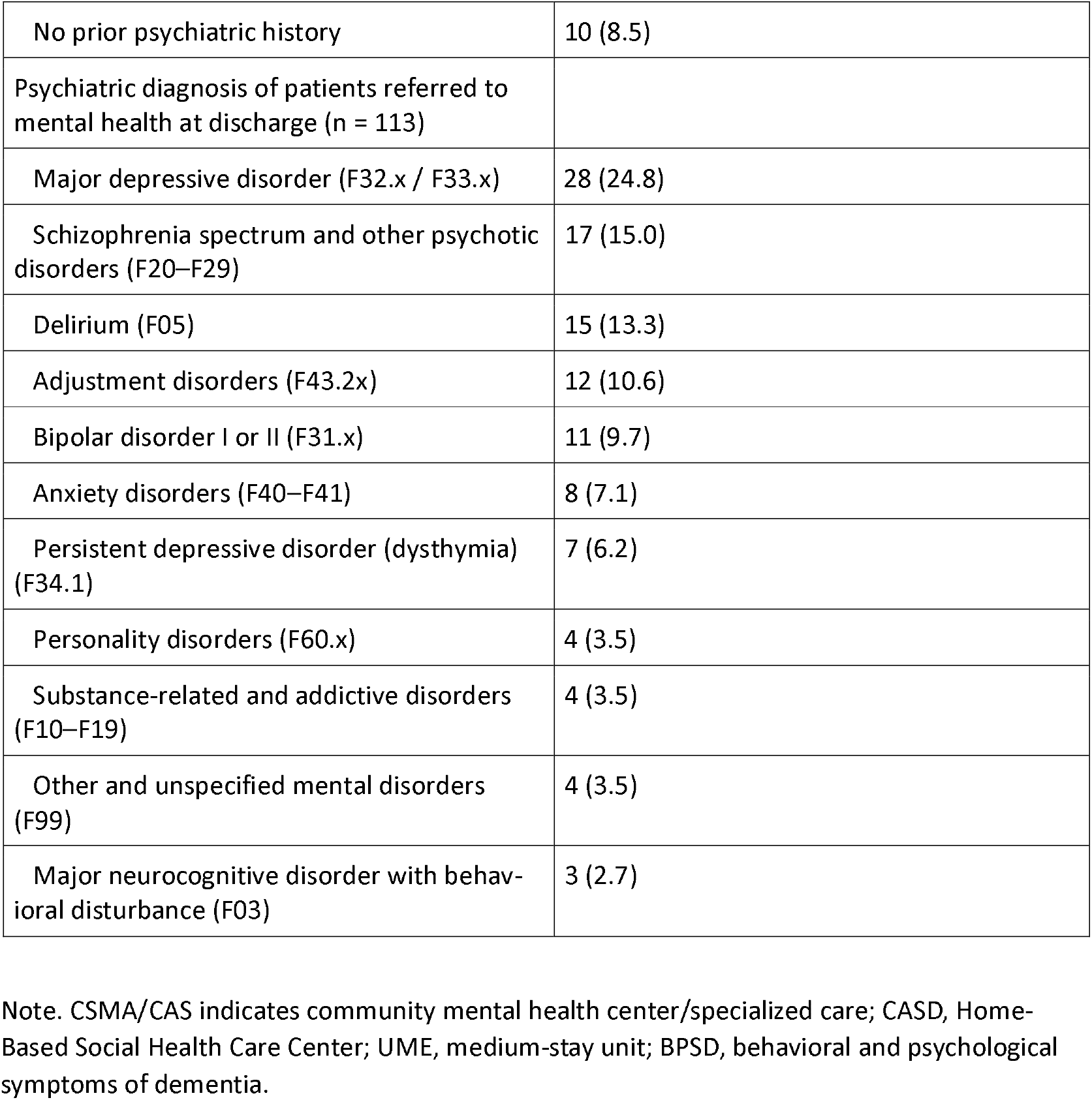
Consultation-Liaison Psychiatry Service Activity, Psychiatric Diagnoses, and Discharge Disposition.

A total of 452 patients with complete information on discharge disposition were analyzed; 25.0% were referred to mental health services at discharge. In the multivariable model, older age was associated with a lower probability of referral (odds ratio [OR] per year, 0.96; 95% CI, 0.93–0.99; p = .005), as was a higher age-adjusted Charlson Comorbidity Index (OR, 0.90; 95% CI, 0.82–1.00; p = .040). Female sex was associated with a higher probability of referral compared with male sex (OR, 1.59; 95% CI, 1.01–2.50; p = .044). The Barthel Index was not associated with referral (OR, 1.00; 95% CI, 0.99–1.01; p = .58).

#### Age Subgroup Analysis

In the cohort (N = 465), 178 patients (38.3%) were aged 65–74 years and 287 (61.7%) were aged ≥75 years. No differences were observed in the Barthel Index by age group (median, 95 vs 90; p = 0.171). In contrast, the ≥75 years group showed lower instrumental functional status (Lawton–Brody Index: median, 4 vs 5; p = 0.000210) and greater frailty (Clinical Frailty Scale: mean, 4.3 vs 3.9; p = 0.012).

Among notable variables, hearing loss was more frequent in patients aged ≥75 years (73.5%) than in those aged 65–74 years (26.5%). Delirium was also more frequently recorded in the ≥75 years group (74.3%) compared with the 65–74 years group (25.7%). Regarding prior follow-up, care in addiction treatment centers or adult community mental health centers (CAS/CSMA) was more frequent among patients aged 65–74 years (29.2%) than among those aged 75 years or older (19.2%)

#### Clinical Events at 1 Month After Discharge: Readmissions and Emergency Department Visits

When analyzing the data from the first month after discharge, valid information was available in 425 patients. Hospital readmissions were recorded in 72 patients (16.9%) and emergency department visits in 102 patients (24.0%).

Among the 71 patients with available information on the reason for hospital readmission, the causes that led to readmission were very heterogeneous medical conditions.

In the 101 patients with available information on the reason for emergency department visits, the most frequent cause was medical-surgical pathology (85.1%), followed by falls (7.9%) and behavioral disturbances (6.9%).

In the multivariable analysis, no statistically significant associations were observed between sex or age and hospital readmission or emergency department visits. A higher Charlson Index was associated with a higher probability of emergency department visits (odds ratio [OR] per point, 1.15; 95% CI, 1.05–1.27), as was greater frailty measured by the Clinical Frailty Scale (OR per point, 1.17; 95% CI, 1.01– 1.35). For the outcome of hospital readmission, no variable reached statistical significance, although a trend was observed with the Charlson Index and frailty.

#### Mortality

In a cohort of 464 patients aged ≥65 years, in-hospital mortality was 5.6%. At 1 month after discharge, among the 432 patients with available follow-up, 21 additional patients died (4.9%). During the period between the first and the third month after discharge, 22 deaths occurred (5.3%).

In the multivariable analysis, detailed in Table 5, age was the only independent predictor of mortality between 1–3 months, whereas greater frailty (CFS) was independently associated with mortality at 1 month.

**Table 5.**
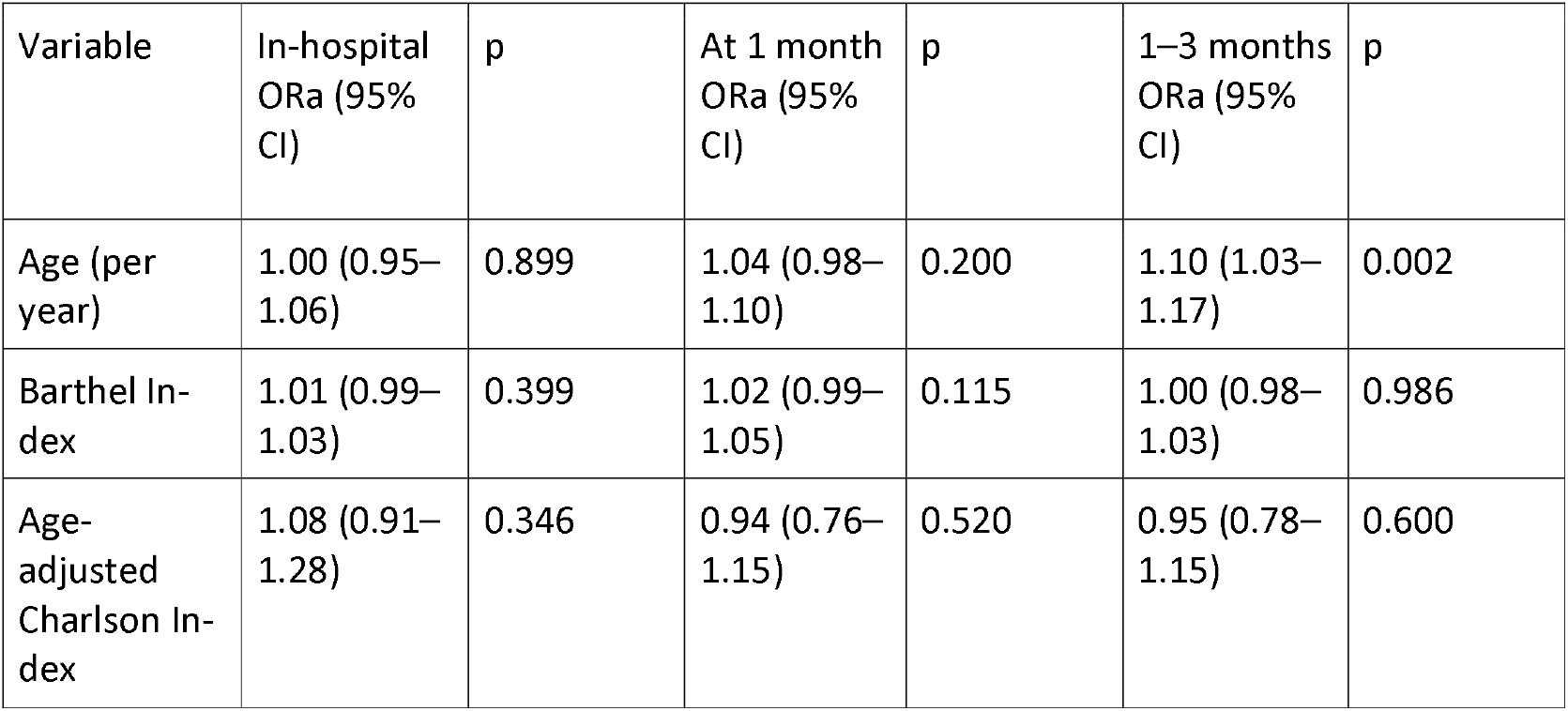
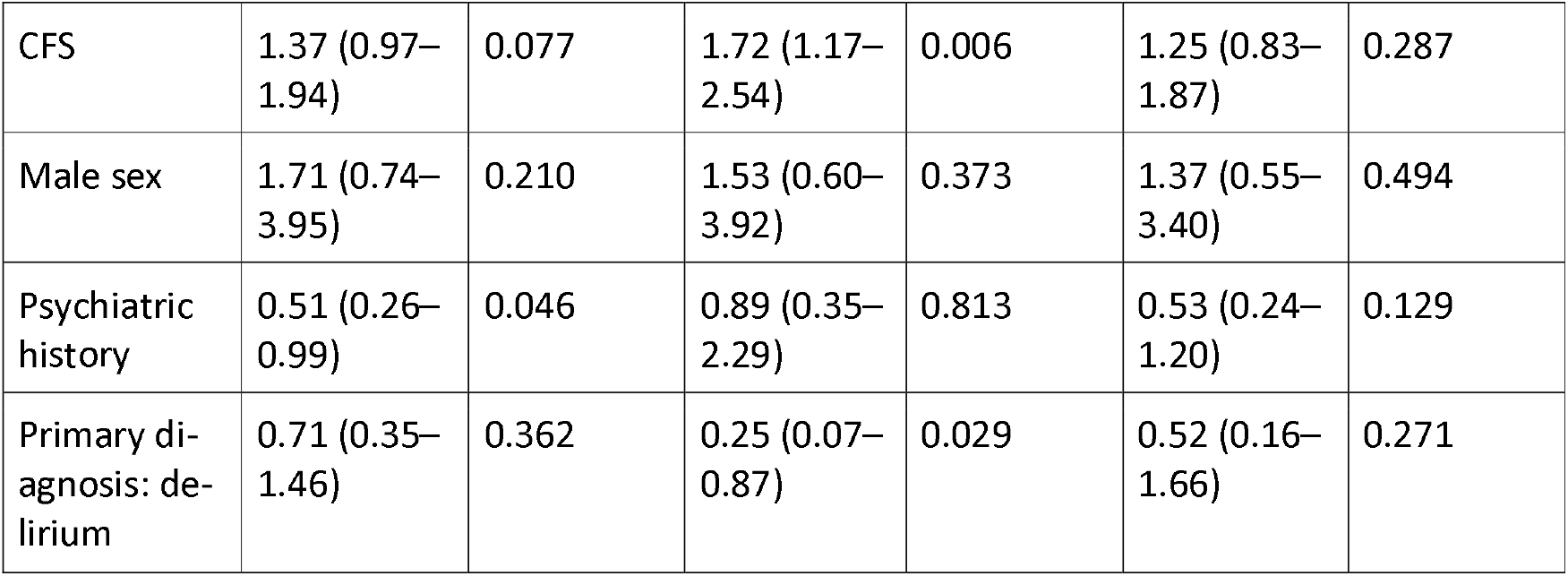
Mortality and Associated Factors (Multivariable Models)

| Variable | In-hospital ORa (95% CI) | p | At 1 month ORa (95% CI) | p | 1–3 months ORa (95% CI) | p |
| --- | --- | --- | --- | --- | --- | --- |
| Age (per year) | 1.00 (0.95–1.06) | 0.899 | 1.04 (0.98–1.10) | 0.200 | 1.10 (1.03–1.17) | 0.002 |
| Barthel Index | 1.01 (0.99–1.03) | 0.399 | 1.02 (0.99–1.05) | 0.115 | 1.00 (0.98–1.03) | 0.986 |
| Age-adjusted Charlson Index | 1.08 (0.91–1.28) | 0.346 | 0.94 (0.76–1.15) | 0.520 | 0.95 (0.78–1.15) | 0.600 |
| CFS | 1.37 (0.97–1.94) | 0.077 | 1.72 (1.17–2.54) | 0.006 | 1.25 (0.83–1.87) | 0.287 |
| Male sex | 1.71 (0.74–3.95) | 0.210 | 1.53 (0.60–3.92) | 0.373 | 1.37 (0.55–3.40) | 0.494 |
| Psychiatric history | 0.51 (0.26–0.99) | 0.046 | 0.89 (0.35–2.29) | 0.813 | 0.53 (0.24–1.20) | 0.129 |
| Primary diagnosis: delirium | 0.71 (0.35–1.46) | 0.362 | 0.25 (0.07–0.87) | 0.029 | 0.52 (0.16–1.66) | 0.271 |

Cumulative mortality at three months was 14.9%, being mainly concentrated in patients with delirium (23.1%) and with adjustment disorder (17.6%).

## DISCUSSION

In this multicenter prospective cohort of older adults evaluated by consultation-liaison psychiatry services (CLPS) across 10 general hospitals, we identified a population characterized by high clinical complexity, substantial medical comorbidity, and a high prevalence of frailty. Clinical factors—particularly frailty and comorbidity—were more strongly associated with outcomes than psychiatric variables alone. These findings highlight the central role of CLPS in the care of hospitalized older adults and support the need for specific training in geriatric psychiatry and the systematic incorporation of comprehensive geriatric assessment into routine clinical practice.

The cohort showed advanced age, a predominance of women, and a socially vulnerable profile, with a considerable proportion of patients living alone or in institutional settings. Although baseline functional status was relatively preserved, hospitalization was associated with a clinically meaningful functional decline, including reduced independence in basic activities and impaired mobility. This pattern is consistent with previous reports in hospitalized older adults and is associated with increased risks of institutionalization, long-term dependency, and mortality, underscoring the complexity of care in this population. ^17^

Frailty was highly prevalent, affecting more than 60% of patients, and was independently associated with early mortality, whereas age emerged as the main predictor of mortality between one and three months after discharge. These findings reinforce the role of frailty as a key prognostic factor in older hospitalized patients and support its systematic use in hospital risk stratification. ^18^

Overall mortality in the cohort was substantial, reaching 14.9% at three months, reflecting the clinical vulnerability of older patients requiring psychiatric consultation during hospitalization. Mortality was particularly frequent among patients with delirium and adjustment disorder, diagnoses that often occur in the context of acute medical illness and underlying frailty. In the adjusted analyses, delirium appeared to be associated with lower mortality at one month. However, this finding should be interpreted cautiously. Because the analysis of one-month mortality included only patients who survived hospitalization, survivor bias may have influenced the observed association. Delirium is known to be associated with increased in-hospital mortality; therefore, patients with delirium who survive to discharge may represent a selected subgroup with greater short-term resilience. In addition, the relatively small number of events may have produced unstable estimates. Consequently, this association should not be interpreted as suggesting a protective effect of delirium. ^19^⍰^20^

Psychiatric morbidity was also substantial, with more than two-thirds of patients having a documented psychiatric history. Delirium remained highly prevalent and clinically relevant, given its association with adverse outcomes and its frequent under recognition. Importantly, more than half of patients received a new psychiatric diagnosis after consultation, highlighting the diagnostic value of CLPS. Even without a structured proactive model, the high rate of newly identified disorders suggests that a significant proportion of psychiatric morbidity remains undetected until specialist assessment. This finding is consistent with previous studies showing that systematic psychiatric evaluation improves detection of previously unrecognized conditions in older adults. ^11^⍰^21^–^23^

Psychotropic medication use was widespread, affecting more than 70% of the cohort, with antidepressants and benzodiazepines being the most frequently prescribed classes. Notably, a substantial proportion of benzodiazepine use occurred in patients without a documented psychiatric history, suggesting potentially inappropriate prescribing in a vulnerable population. The high use of quetiapine likely reflects its off-label use as a hypnotic despite limited evidence and safety concerns in older adults. These findings are consistent with recent studies reporting increasing psychotropic use in older populations, particularly in the context of multimorbidity and polypharmacy. ^24^⍰^25^

Psychotropic polypharmacy was common and was associated with a higher prevalence of falls, supporting evidence linking cumulative medication burden with fall risk in older adults. ^26^–^28^ Although pregabalin and gabapentin use was more frequent among patients with falls, this association did not reach statistical significance, possibly because of limited statistical power or confounding by indication. Nevertheless, both drugs are recognized as fall-risk–increasing medications, and pharmacovigilance data support their association with fall-related adverse events. ^26^–^29^ No significant differences were observed for benzodiazepines or antipsychotics. The risk attributed to these medications may be influenced by the underlying clinical indication and by periods of clinical instability preceding treatment initiation, complicating direct causal attribution. ^20^⍰^31^ Overall, these findings support structured medication review as a priority intervention in older adults with a history of falls and highlight the potential role of CLPS in deprescribing strategies.^27^⍰^32^

We also observed an inverse association between age and prior access to specialized mental health services among patients with psychiatric history. After adjustment for clinical and functional variables, age remained the only factor independently associated with lower likelihood of specialized follow-up. This finding suggests that disparities in access to mental health care among older adults may not be fully explained by clinical burden alone and are consistent with previous literature describing structural barriers and age-related inequalities in mental health care access. ^32^–^34^

The lower probability of referral to mental health services at discharge among older patients appeared to be largely explained by clinical and functional factors. Older patients were more frequently referred to social-health resources or post-acute care settings, indicating that discharge planning was primarily driven by dependency and care needs rather than psychiatric diagnosis. This interpretation is supported by previous studies showing that functional status and frailty are key determinants of discharge planning and post-acute care utilization. ^35^⍰^36^

Age-stratified analyses further supported the existence of distinct clinical profiles. Older patients showed greater functional dependence, higher frailty, lower substance use, and different diagnostic patterns. These findings reinforce the need for age-sensitive and individualized approaches to care in consultation-liaison psychiatry. ^5^⍰^9^⍰^37^⍰^38^

This study has several limitations. Psychiatric diagnoses were based on routine clinical assessment without structured diagnostic interviews, which may have introduced interobserver variability. Pharmacological data were limited to the presence or absence of treatment prior to admission, without information on dosage, duration, or adherence. In addition, patients unable to provide informed consent and without a legal representative were excluded, potentially underrepresenting the most severe clinical profiles. Finally, the observational design precludes causal inference despite multivariable adjustment.

Overall, these findings identify patients evaluated by CLPS as a high-risk population in whom frailty, psychiatric morbidity, and high exposure to psychotropic medications converge. These findings have potential implications for clinical practice. Systematic assessment of frailty and functional status may help identify older hospitalized patients at increased risk of adverse outcomes following psychiatric consultation. In this context, consultation-liaison psychiatry services may represent a key setting for integrating geriatric assessment, optimizing psychotropic prescribing, and coordinating multidisciplinary care for medically complex older adults during hospitalization.

## Data Availability

All data produced in the present study are available upon reasonable request to the authors

## Conflict of Interest Disclosures

The authors declare no conflicts of interest.

## Funding/Support

This study received no external funding.

## Supplement 1. Participating Hospitals

Hospital Universitario La Princesa; Hospital Universitario del Sureste; Hospital Clínico San Carlos; Hospital Universitari Germans Trias i Pujol; Hospital Clinic i Provincial; Hospital de la Santa Creu i Sant Pau; Hospital Vall d’Hebron; Hospital Universitario de Bellvitge; Corporació Sanitaria Parc Taulí; Consorci Sanitari Integral.

## Notes

### Competing Interest Statement

The authors have declared no competing interest.

### Author Declarations

The study was approved by the Research Ethics Committee of Hospital Universitari de Bellvitge (Ref. PR069/23 [CSI 23/13]) and by the appropriate regulatory au-thorities at all participating centers

